# Predicting hearing help-seeking: What features are important for a profiling module within a hearing mHealth application?

**DOI:** 10.1101/2023.02.14.23285904

**Authors:** Giulia Angonese, Mareike Buhl, Inka Kuhlmann, Birger Kollmeier, Andrea Hildebrandt

## Abstract

**Background:** Mobile health care solutions can improve quality, accessibility and equity of health services, fostering early rehabilitation. For people suffering from hearing loss, mobile applications might be designed to support the decision-making processes in auditory diagnostics and to provide treatment recommendations to the user (e.g., hearing aid need). For some individuals, such mobile app might be the first contact with a hearing diagnostic service and should motivate users with hearing loss to seek professional help.

**Objective:** This study aims at characterizing individuals who are more or less prone to seek professional help after the repeated use of an app-based hearing test. The goal is to develop a profiling module building upon hearing related traits and personality characteristics to secure personalized treatment recommendations in hearing mHealth solutions.

**Methods:** *N*=185 (106 females) non-aided older individuals (*M*_age_=63.8, *SD*_age_=6.6) with subjective hearing loss participated in a comprehensive online study. We collected cross-sectional and longitudinal data on several hearing-related and psychological features that were previously found to predict hearing help-seeking. Readiness to seek help was assessed as outcome variable at study-end and after two months. Participants were classified into help-seekers and non-seekers with several supervised machine learning algorithms (Random Forest, Naïve Bayes and Support Vector Machine). The most relevant features for prediction were identified with feature importance analysis.

**Results:** The algorithms correctly predicted action to seek help at study-end in 66 to 70% of cases, reaching 75% classification accuracy at follow-up. Among the most important features for classifications were the degree of hearing loss and its perceived consequences in daily life, attitude towards hearing aids, and the personality traits neuroticism and conscientiousness.

**Conclusions:** This study contributes to the identification of individual characteristics that predict help-seeking in older individuals with self-perceived hearing loss. Suggestions for the implementation of an individual profiling algorithm and for targeted recommendations in hearing mHealth applications are derived.

## Introduction

### Mobile health solutions for hearing care

Hearing enables people to experience their surroundings and to communicate with others. Thus, hearing difficulties can have a strong impact on individuals’ quality of life. Hearing loss (HL) is one of the most common chronic diseases worldwide and it affects 20.3% of the world’s population. More than 60% of individuals with HL are older than 50 years of age, with the principal cause being age-related HL [1]. Untreated hearing difficulties have been associated with lower self-rated health, depression, and anxiety, in addition to physical and cognitive decline, dementia, and hospitalization in the older population [2– 4]. The primary rehabilitative strategy for individuals with moderate to severe HL is the use of hearing aids (HA), which increases activity levels, general health, quality of life [5] and decreases social isolation and depressive symptoms [6] by supporting hearing ability and communication efficacy. Despite the positive effects of hearing rehabilitation, the prevalence of HA use is still limited to about 25% of the hearing impaired population [2,4,7,8]. Moreover, there is an average delay of nine years between the time a person first acknowledges hearing difficulties until the first contact with a hearing-health professional [9].

Developing easily accessible and affordable mobile health (mHealth) solutions in audiology could promote broader and faster access to diagnosis and health services, fostering an early rehabilitation and reducing the impact of HL on the individual. Before such mHealth tools are implemented, there is need for basic research on potential components of an app that could provide audiological self-test options and HA algorithms for self-fitting, both with individualized recommendations. Through such an app, users could receive specific treatment recommendations depending on their hearing performance, including suggestions to visit a hearing acoustician or ENT (Ear, Nose and Throat) physician, if indicated. In addition, users could be given the possibility to experience listening conditions of a HA and its potential benefits. However, even though professional support might be recommended, a fair amount of users might still be reluctant to seek help and need more convincing incentives in order to take action. A hearing mHealth app might often represent the first contact with a hearing diagnostic service and should therefore motivate individuals in need (especially the predicted “non-help-seekers”) to pursue professional help, in order to optimize the impact on the population. It follows that the assessment of an individual with hearing difficulties should go beyond simply quantifying HL and should consider individual characteristics that have been shown to influence the readiness of individuals to seek professional help [4,10]. Moreover, such information collected by a hearing mHealth app could later help clinicians in performing a personalized counseling.

### Predictors of hearing help-seeking and hearing aid uptake

Hearing help-seeking can be seen as a first crucial step towards the decision to uptake a HA [10]. Help-seeking takes place after a contemplation stage [3], where a listener in need is initially ambivalent about making changes. Help-seekers would then prepare (seek information, plan) and take action [3] towards a change in behavior and attitude, namely consulting a healthcare professional about their hearing difficulties [4,11]. Acknowledgement and acceptance of hearing difficulties and their impact on everyday life have been discussed as the most important predictive features of hearing help-seeking, as well as later HA uptake [5,6,10,12,13]. HL is frequently perceived as part of the natural ageing process and other health issues are prioritized for treatment [4,14]. Even when HL is identified, individuals might reject the use of a HA due to expected costs, stigma and negative stereotypes [5,7,12]. However, a positive attitude towards HA [10,11], high expectations to benefit from them [10,13] and perceived self-efficacy in their daily management [14,15] were shown to promote help-seeking and HA uptake. Further relevant predictors are personal attitudes, beliefs, and personality traits. Individuals who are more prone to seek help and successfully uptake a HA show higher internal locus of control [2, 14] self-efficacy [15,16], and agreeableness, as well as lower neuroticism and openness [2]. Altogether such individual characteristics refer to a general self-confidence in the ability to cope with critical situations, good acceptance of others’ suggestions and recommendations, as well as less susceptibility to shame and embarrassment and less analytical thinking.

### Rationale and objective

The present study aims at identifying the most important predictive features for hearing help-seeking, aiming to design an individual profiling module for a hearing mHealth app. Such module will categorize help-seekers versus non-seekers and ultimately inform the design of targeted or even personalized treatment recommendations. For this purpose, data from a large number of questionnaires and tests related to different hearing-related and psychological characteristics, together with multiple assessments of a hearing screening test were collected in a mobile study that simulated a hearing mHealth app. To target potential users of future apps of this kind, the study was addressed to individuals with subjectively perceived hearing difficulties who had not yet been compensated with HA. We used supervised machine learning algorithms to predict the readiness to seek professional help, as assessed at the end of the study and after two months by means of features that are characteristic of a typical hearing help-seeking population.

The following research questions guided our study design and analyses:

RQ1. Which machine learning model can best predict help-seeking and categorize individuals into help-seekers versus non-seekers?
RQ2. Which hearing-related and psychological features are most relevant to profile individuals and tell help-seekers versus non-seekers apart?
RQ3. How can feature importance measures inform the design of targeted recommendations for users of a future hearing mHealth app?

## Methods

### Study overview

The study was conducted entirely online on personal mobile phones of the participants to approximate the experience of dealing with a mobile app. Communications with the participants took place via email and SMS. The total assessment time of eight hours was distributed across the working days of three consecutive weeks, with an overall daily testing time of approximately 30 minutes. The first week (baseline assessment) included one measurement per day, which could be performed at any preferred time. The second and third week included two measurement time-points per day (morning and evening) of approximately 15 minutes each. The data from this longitudinal assessment were summarized for subsequent analysis, as described below (see Statistical analysis section). Participants were remunerated with 10 Euros per hour. After two months from the end of the study, participants were invited to complete a voluntary follow-up online questionnaire.

### Ethics approval and data availability

The study plan and data management have been approved by the Research Ethics Committee of the Carl von Ossietzky Universität Oldenburg. We, as single researchers and as a team, are committed to and supporting Open Science practices. We have therefore published a preprint of the manuscript on medrxiv.org and share data analysis scripts via Zenodo (please refer to the data availability statement for the links). The data will be shared with interested researchers upon request, since the dataset will be used for further projects and cannot be fully shared at the current time point.

### Participants

Older adults above 50 years of age were recruited between September 2021 and August 2022 through the online platform Ebay’s minijob announcements, the university intranet and via mailing list services of several German universities’ Guest-Audience and Senior-University programs. Inclusion criteria were: subjective reports of hearing difficulties in daily life, ownership of and the ability to use a smartphone, and good command of the German language. The exclusion criterion was the use of hearing aids. Prior to enrolment, participants provided written informed consent. The final dataset included *N*_1_ = 185 participants, 106 female and 79 male (0 diverse), aged between 47 and 82 years, with *M*_age_ = 63.1 and *SD*_age_ = 6.5. One participant was below 50 years of age (47) but was nevertheless included in the final sample, given that this value only slightly deviated from the planned age threshold. Out of the *N*_1_=185 participants who completed the study, *N*_2_=131 attended the follow-up questionnaire. A descriptive summary of participants’ socio-demographic characteristics is provided in Table 1.

**Table 1.** Main socio-demographic characteristics of the participants^a^.

| Characteristic |  | *n* (% of total) |
| --- | --- | --- |
| Age group (years) |  |  |
|  | 47-60 | 70 (37.8) |
|  | 61-70 | 83 (44.9) |
|  | 70-82 | 32 (17.3) |
| Sex |  |  |
|  | female | 106 (57.3) |
| Duration of hearing difficulties (years) |  |  |
|  | 0 - 1 | 54 (29.2) |
|  | 2 - 5 | 89 (48.1) |
|  | 6 - 10 | 23 (12.4) |
|  | 10 – 21 | 19 (10.3) |
| Presence of Tinnitus |  |  |
|  |  | 65 (35.1) |
| Previous doctor consultation for hearing difficulties |  |  |
|  |  | 89 (48.1) |
| Presence of visual problems |  |  |
|  |  | 134 (72.4) |
| Occupation status |  |  |
|  | employed | 67 (36.2) |
| Monthly income (Euros) |  |  |
|  | < 1.500 | 62 (33.5) |
|  | 1.500 - 2.500 | 65 (35.1) |
|  | 2.500 – 4.000 | 42 (22.7) |
|  | >4.000 | 16 (8.7) |
| Residential environment |  |  |
|  | countryside | 2 (1.1) |
|  | little town | 29 (15.7) |
|  | suburbs | 48 (25.9) |
|  | city | 106 (57.3) |
| Self-estimated noise level at home |  |  |
|  | low | 67 (36.2) |
|  | moderate | 113 (61.1) |
|  | high | 5 (2.7) |

<sup>a</sup> These data were acquired during the baseline assessment through a self-report questionnaire.

### Procedure

#### Online tools

The data collection was carried out with the online, open-source software formr [17]. Here, different questionnaires and tests (see section Assessment) were combined as a survey in a run that reproduced the desired design and could be assessed by users through a specific link. The Customer Communication Platform Twilio [18] was used to send the study link to the participants through daily text message reminders. With the individualized link received via SMS, participants could perform the study on their own smartphone’s browser. Seven participants performed the study on their computers since they experienced technical difficulties with their smartphones.

#### Baseline assessment

During baseline assessment, cross-sectional data from a comprehensive set of 25 questionnaires and tests was collected. The questionnaires and tests were distributed on five consecutive days in order to maximise study compliance and avoid priming effects on different questionnaires. For a detailed list of all assessment tools and their references, as well as the derived features for analysis, we refer to Table 2 and Table 3. The following is a brief overview of the predictors assessed.

**Table 2.**
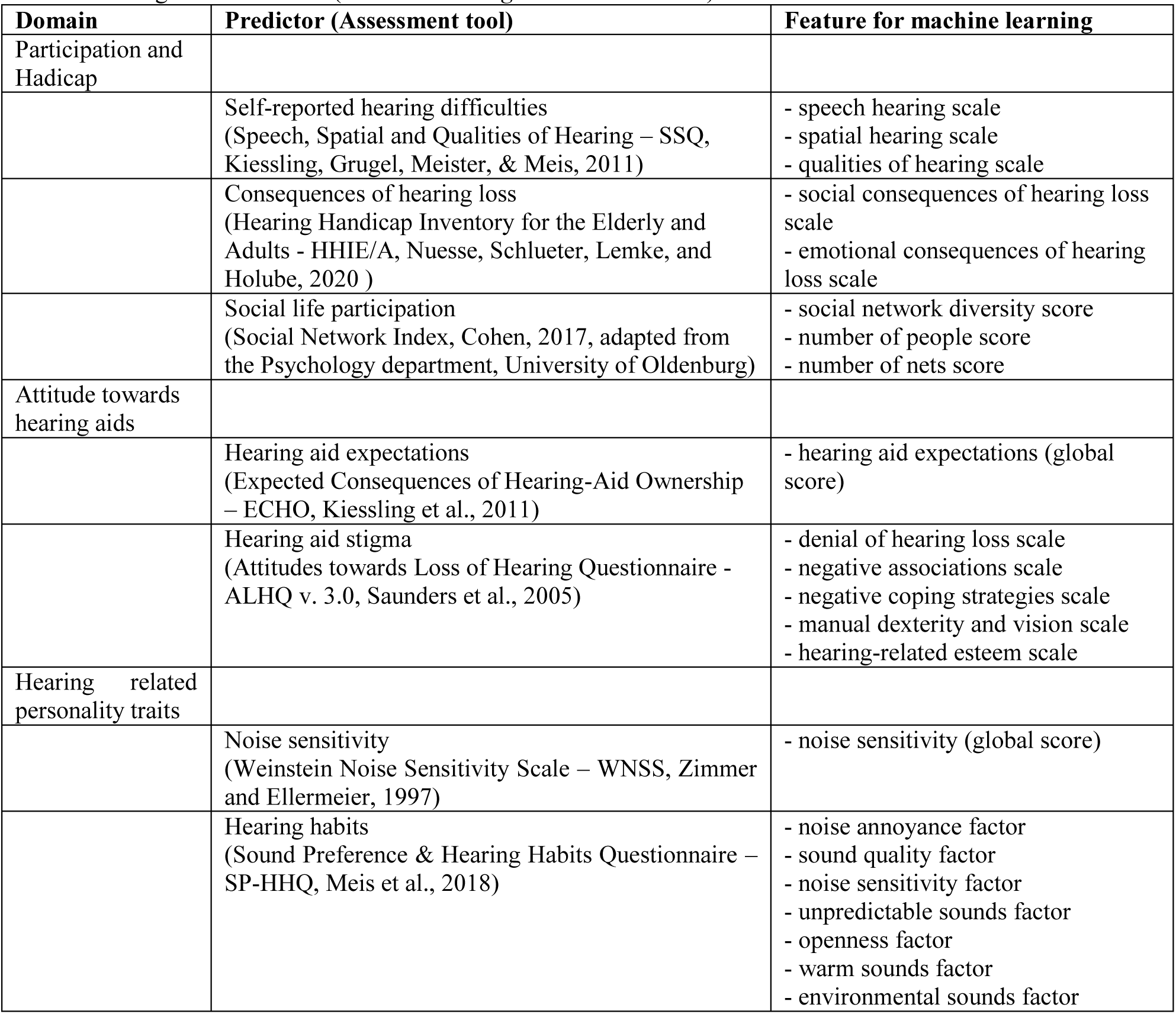
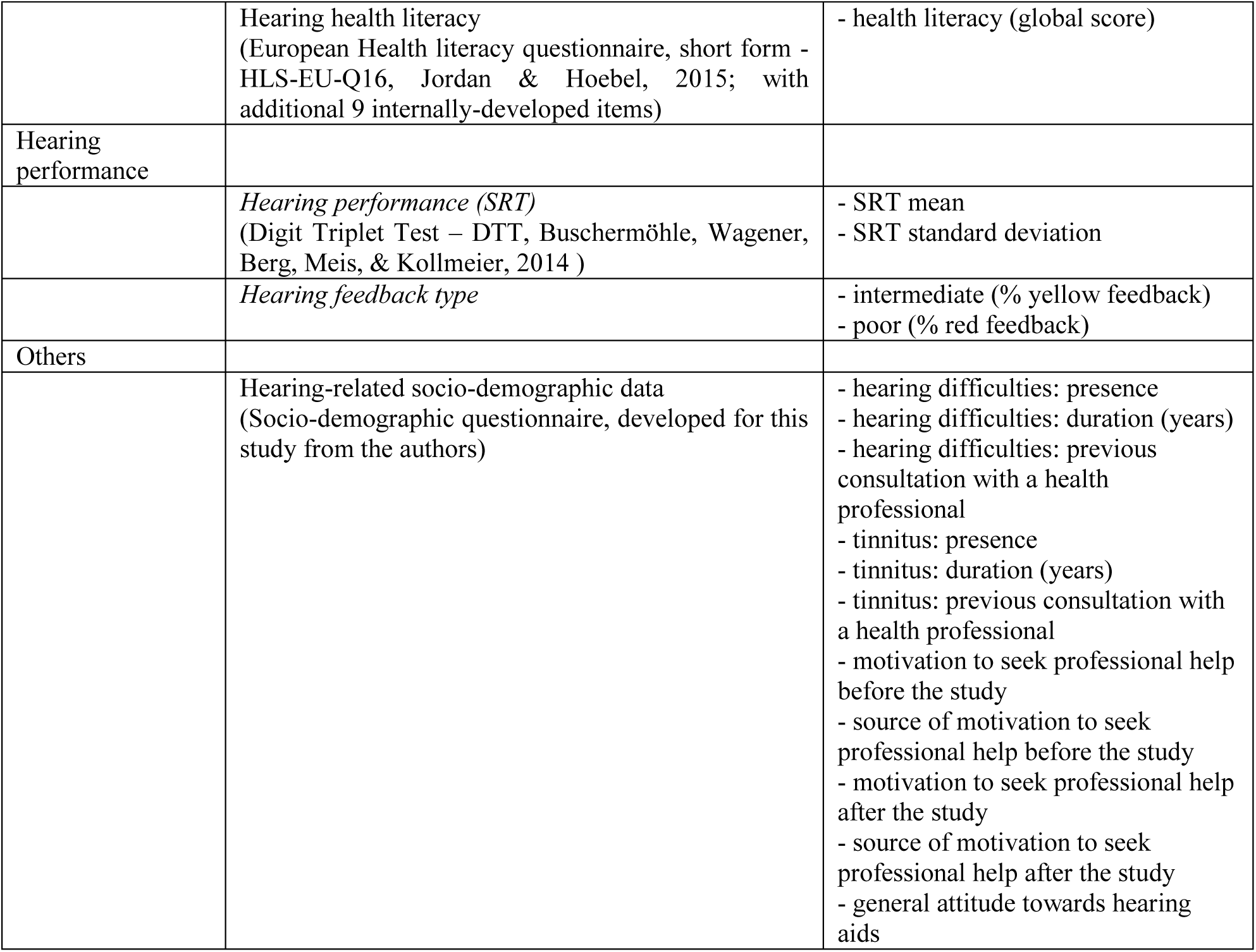
Hearing-related features (baseline and *longitudinal* assessment).

| Domain | Predictor (Assessment tool) | Feature for machine learning |
| --- | --- | --- |
| Participation and<br>Hadicap |  |  |
|  | Self-reported hearing difficulties<br>(Speech, Spatial and Qualities of Hearing – SSQ, Kiessling, Grugel, Meister, & Meis, 2011) | - speech hearing scale<br>- spatial hearing scale<br>- qualities of hearing scale |
|  | Consequences of hearing loss<br>(Hearing Handicap Inventory for the Elderly and Adults - HHIE/A, Nuesse, Schlueter, Lemke, and Holube, 2020 ) | - social consequences of hearing loss scale<br>- emotional consequences of hearing loss scale |
|  | Social life participation<br>(Social Network Index, Cohen, 2017, adapted from the Psychology department, University of Oldenburg) | - social network diversity score<br>- number of people score<br>- number of nets score |
| Attitude towards<br>hearing aids |  |  |
|  | Hearing aid expectations<br>(Expected Consequences of Hearing-Aid Ownership – ECHO, Kiessling et al., 2011) | - hearing aid expectations (global score) |
|  | Hearing aid stigma<br>(Attitudes towards Loss of Hearing Questionnaire - ALHQ v. 3.0, Saunders et al., 2005) | - denial of hearing loss scale<br>- negative associations scale<br>- negative coping strategies scale<br>- manual dexterity and vision scale<br>- hearing-related esteem scale |
| Hearing related<br>personality traits |  |  |
|  | Noise sensitivity<br>(Weinstein Noise Sensitivity Scale – WNSS, Zimmer and Ellermeier, 1997) | - noise sensitivity (global score) |
|  | Hearing habits<br>(Sound Preference & Hearing Habits Questionnaire – SP-HHQ, Meis et al., 2018) | - noise annoyance factor<br>- sound quality factor<br>- noise sensitivity factor<br>- unpredictable sounds factor<br>- openness factor<br>- warm sounds factor<br>- environmental sounds factor |
|  | Hearing health literacy<br>(European Health literacy questionnaire, short form - HLS-EU-Q16, Jordan & Hoebel, 2015; with additional 9 internally-developed items) | - health literacy (global score) |
| Hearing performance |  |  |
|  | *Hearing performance (SRT)*<br>(Digit Triplet Test – DTT, Buschermöhle, Wagener, Berg, Meis, & Kollmeier, 2014 ) | - SRT mean<br>- SRT standard deviation |
|  | *Hearing feedback type* | - intermediate (% yellow feedback)<br>- poor (% red feedback) |
| Others |  |  |
|  | Hearing-related socio-demographic data<br>(Socio-demographic questionnaire, developed for this study from the authors) | - hearing difficulties: presence<br>- hearing difficulties: duration (years)<br>- hearing difficulties: previous consultation with a health professional<br>- tinnitus: presence<br>- tinnitus: duration (years)<br>- tinnitus: previous consultation with a health professional<br>- motivation to seek professional help before the study<br>- source of motivation to seek professional help before the study<br>- motivation to seek professional help after the study<br>- source of motivation to seek professional help after the study<br>- general attitude towards hearing aids |

**Table 3.** Assessment of psychological, general health and socio-demographic features (baseline and longitudinal assessment).

| Domain | Predictor (Assessment tool) | Feature for machine learning |
| --- | --- | --- |
| Personality traits |  |  |
|  | Big 5<br>(NEO-ffi, Borkenau & Ostendorf, 1993) | - neuroticism scale<br>- extraversion scale<br>- openness scale<br>- agreeableness scale<br>- conscientiousness scale |
|  | Trait anxiety<br>(Geriatric Anxiety Inventory, Gottschling, Segal, Häusele, Spinath, & Stoll, 2016) | - anxiety (global score) |
|  | Trait depression<br>(Geriatric Depression Scale, Yesavage & Sheikh, 1986) | - depression (global score) |
|  | Optimism/Pessimism<br>(Die Skala Optimismus-Pessimismus – 2, Kemper, Beierlein, Kovaleva, & Rammstedt, 2012) | - optimism (global score) |
|  | Loneliness<br>(De Jong Gierveld Loneliness Scale - DJG scale, Tesch-Römer et al., 2013) | - loneliness (global score) |
|  | Sensory processing sensitivity (High Sensitive Person Scale - HSPS-G, Konrad and Herzberg, 2019) | - ease of excitation scale<br>- sensory threshold scale<br>- aesthetic sensitivity scale<br>- hearing scale |
| Attitudes and beliefs |  |  |
|  | Health Locus of Control<br>(Kontrollüberzeugungen zu Krankheit und Gesundheit – KKG, Lohaus & Schmitt, 1989)<br>(Internale-Externale Kontrollüberzeugung-4 Skala, Kovaleva et al., 2012) | - internal locus of control scale<br>- society control scale<br>- external locus of control scale |
|  | Attitude toward ageing<br>(Attitude to ageing questionnaire – AAQ, Laidlaw, Power, & Schmidt, 2007) | - psychosocial scale<br>- physical scale<br>- psychological scale |
|  | General self-efficacy<br>(Generalized Self-Efficacy Scale – GSES, Schwarzer & Jerusalem, 2003) | - general self-efficacy (global score) |
| Mood |  |  |
|  | *Affect*<br>(Daily questionnaire on affect, developed for this study from the authors) | - positive affect pre-test mean<br>- positive affect pre-test standard deviation<br>- negative affect pre-test mean<br>- negative affect pre-test standard deviation<br>- positive affect post-test mean<br>- positive affect post-test standard deviation<br>- negative affect post-test mean<br>- negative affect post-test standard deviation |
|  | Stress<br>(Perceived Stress Scale – PSS, E. Schneider, Schönfelder, Domke-Wolf, & Wessa, 2020) | - perceived stress (global score) |
| Cognitive functions |  |  |
|  | Figural Reasoning<br>(Berlin Fluid and Crystallized Intelligence Task – BEFKI, Schipolowski, Wilhelm, & Schroeders, 2020) | - figural reasoning (global score) |
|  | Vocabulary<br>(Wortschatztest, Schmidt & Metzler, 1992) | - vocabulary (global score) |
|  | Digital literacy<br>(Technikbereitschaft -Kurzskala, Neyer et al., 2012) | - technology commitment (global score) |
| Others |  |  |
|  | General health<br>(Fragebogen zum Allgemein Gesundheitszustand SF-12, Drixler, Morfeld, Glaesmer, Brähler, & Wirtz, 2020 ) | - physical health score<br>- mental health score |
|  | General Socio-demographic data<br>(Socio-demographic questionnaire, developed for this study from the authors) | - age<br>- sex<br>- presence of visual problems<br>- education degree<br>- occupation (retired or working)<br>- weekly working hours<br>- monthly income |
|  |  | - relationship status<br>- monthly income of the partner<br>- residential environment<br>- household size |

##### Assessment of hearing-related features

Firstly, the assessment included self-reports of participation and perceived handicap, focusing on self-reported hearing difficulties, consequences of HL and social life participation. Additionally, attitudes towards HA were evaluated with questionnaires on HA expectations and stigma. Hearing-related personality traits were also taken into account. Noise sensitivity was measured as a personality trait which was shown to be related to affect and neurosensory processing [19]. We further assessed hearing habits, aiming to gather more information about sound sensitivity and individuals’ sound preference profiles [20]. Finally, hearing health literacy was assessed as well, since the ability to search, find and understand information related to hearing health was shown to be associated with better self-management of HL [15].

##### Assessment of psychological, general health and socio-demographic features

Personality traits (the Big Five [21]) were shown to be associated with help-seeking and HA use, and were therefore included in the baseline assessment. Anxiety and depression were also measured, given their frequently demonstrated associations with HL [4,6], together with loneliness, which is seen as consequence of untreated HL [3]. We further assessed optimism and sensory processing sensitivity, which refers to an individual’s disposition to perceive and process stimuli (including auditory ones) more intensely than the average population [22]. Attitudes and beliefs like locus of control and self-efficacy were included as well for their association with help-seeking and HA use. The belief that HA are associated with old age and infirmity is often a barrier to HA uptake and use [4], therefore attitude towards ageing was assessed as well. Perceived stress was measured too, since high levels of stress that are related to daily life, work or social situations may boost help-seeking behaviours [3]. General health was assessed given its predictive role for different steps of the HA uptake path [4,6]. For completeness, we also measured cognitive abilities (crystallized and fluid intelligence), despite discordant findings on associations between cognition and HA uptake [4,6,13]. Lastly, participants were requested to complete a comprehensive questionnaire on socio-demographics.

#### Longitudinal assessment of hearing and affect

This micro-longitudinal assessment accounts for potential daily fluctuations of hearing performance and affect, which might depend on particular daily events and states. The affect questionnaire included 14 items in line with the Circumplex Model of Affect (Russel, 1980 [24]). Eight items were related to negative affect and six to positive affect [25]. The items are listed in the Multimedia Appendix 1. The affect questionnaire was presented before and after the hearing test in order to assess mood at baseline and after receiving feedback on the hearing test, respectively.

Hearing performance was assessed with the Digit Triplets Test (DTT) [26,27] of the Hörzentrum Oldenburg gGmbH. This screening instrument measures speech intelligibility in noise by means of the Speech Recognition Threshold (SRT), which indicates the Signal-to-Noise Ratio (SNR, difference between speech and noise level) at which the participant reaches 50% speech intelligibility. After completing the hearing test, participants received a feedback on their performance in the form of a traffic-light colour, where green indicated good (SRT < −7.1 dB SNR), yellow indicated intermediate (−7.1 >= SRT < −5.1 dB SNR), and red reflected poor performance (SRT >= −5.1 dB SNR) [27,28]. The appendix provides detailed information on the hearing test (Multimedia Appendix 1) and its feedback (Multimedia Appendix 2).

#### Outcome measures

At the end of the study, participants were asked to report on planned actions to seek professional help for their perceived hearing difficulties and their motivation to seek help. This information was also assessed at the beginning of the study. These variables, as retrieved at the end of the study, were chosen as outcome measures for the supervised machine learning (see below). The distribution of participants along the outcome classes considered is summarized in Table 4.

**Table 4.** Absolute class-wise frequencies of observations at 2 month follow-up across the three outcomes considered.^a^.

|  | **action**<br>**at follow-up** | **no action**<br>**at follow-up** | **no follow-up data** | total |
| --- | --- | --- | --- | --- |
| **action** | 33 | 12 | 19 | 64 |
| **no action & high**<br>**motivation** | 12 | 22 | 13 | 47 |
| **no action & low**<br>**motivation** | 7 | 45 | 22 | 74 |
| total | 52 | 79 | 54 |  |

<sup>a</sup> The three outcomes considered are: action to seek help ( $N_1=185$ ); action and motivation to seek help ( $N_1=185$ ); action to seek help at follow-up ( $N_2=131$ ). Additionally, the table provides an overview of those participants who did not complete the follow-up questionnaire.

### Action to seek help

Help-seeking (preparation and action [3]) was assessed at study end with the question: “Given the feedbacks of this study regarding your hearing performance, have you made an appointment with one of the following doctors or a hearing care professional, or are you planning to do so?” (followed by a list of hearing professionals). This variable was used to create two outcome classes:

▪ *action* class (*n_11_*= 64): participants who were planning to seek professional help in the near future or had already taken an appointment;
▪ *no action* class (*n_12_* = 121): participants not ready to take action, who did not plan to consult a hearing health professional in the future.

### Action and motivation to seek help

A second outcome measure was taken into account to further differentiate the *no action* class, in order to provide further insight for the design of targeted recommendations in a hearing mHealth app. Information on readiness to take action was combined with the reported motivation to seek help at the end of the study. Motivation was assessed through the question: “How motivated are you at the moment to seek help regarding your hearing problems?” (1 = not motivated at all, 7 = very strongly motivated). The answer spectrum was binarized by means of median split to create the following outcome classes:

▪ *no action & high motivation* class (*n_13_* = 47): participants not ready to take action with high motivation, who might particularly benefit from personalised and tailored recommendations;
▪ *no action & low motivation* class (*n_14_* = 74): participants not ready to take action who reported low motivation to seek help;
▪ *action* class (*n_15_* = 64): participants ready to take action, regardless of their motivation level. This class was not further divided with respect to motivation, since this would not result in different recommendations. Moreover, data exploration revealed that only seven individuals out of 64 in this category reported low motivation.

### Action to seek help at a follow-up

Two months after the end of the study, participants who agreed to be contacted for a follow-up received a further questionnaire (in formr). The short survey consisted of two multiple-choice questions and it was completed by 71% (*N_2_* = 131) of the participants. Individuals were asked to report again on their action to seek professional help. In addition, they were asked to indicate whether the study participation improved their awareness towards hearing difficulties. Only the first question was considered as further outcome measure. The answers (given as four multiple-choice answers) were binarized to achieve a class allocation comparable to the first outcome measure:

▪ *action at follow-up* class (*n_21_* = 52): participants who reported to have completed an appointment with a hearing professional, who had an appointment fixed, yet not completed or who were planning to seek help in the near future;

▪ *no action at follow-up* class (*n_22_* = 79): participants who did not plan to consult a hearing health professional;

### Statistical analysis

#### Data pre-processing

Data analysis was performed with the R Software for Statistical Computing [29]. Raw data from all questionnaires was imported from the online platform formr to R environment using the dedicated package formr [30]. For each questionnaire or test presented at baseline, global scores were computed and considered as features. If both global scores and scale scores were available for a given assessment tool, only the scale scores were maintained if considered differentially relevant for the outcome. Hearing test results were received from the Hörzentrum Oldenburg and imported in R as .eml files. The performance feedback category (green, yellow, red) was additionally extracted and stored for each raw SRT result. Due to the particular implementation of the study in formr, participants could perform the hearing-test more than once at each measurement time-point. Whenever this happened, only the last SRT result at a given time-point was kept for analysis. This led to a removal of 3.9% of the raw SRT results. The longitudinal data on daily mood and hearing performance was summarized into individual means and standard deviations. The summarized longitudinal data was merged with the cross-sectional data, resulting in a wide-format data frame including 88 features. Table 2 and 3 provide a complete list of features considered for analysis.

Missing data occurred for the longitudinal measures of hearing performance and affect. A complete set of 20 SRT results was collected for 43.8% of participants, while at least 15 SRT results were obtained in 95.1% of cases. Missing hearing data at a specific time-point was considered Not Available (NA). Where an SRT result was missing, the respective feedback and measures of affect at post-test were set to missing as well (NA). By visual inspection of the individual SRT distributions, some specific outlier patterns were identified. 14 participants showed a much larger SRT result at the first measurement, which qualified as outlier following the interquartile range rule. These large SRT values (indicating poor performance) were considered to be caused by misunderstanding of the hearing test instructions and were therefore set to NA. The respective feedback category and measures of affect where however not set to NA. This is because despite the unreliable SRT value, participant’s mood could still have been affected by the feedback received. With respect to daily affect measures, two participants provided no data at measures of post-test affect, such that summary measures could not be computed. In these cases, mean imputation technique was applied: the sample mean and sample variance for negative and positive affect at post-test were imputed to replace missing values.

#### Machine learning

The data was fed into three machine learning algorithms for supervised classification. We chose Naïve Bayes (NB), Random Forests (RF) and Support Vector Machines (SVM) among other classifiers to cover a wide range of model complexity (from simple models like NB, to more complex and non-linear ones like SVM). The algorithms have been implemented on R using the mlr package [31], following the approaches described in [32] and [33]. Given the presence of three different outcome measures, the same analysis steps were carried out in parallel for each outcome, with a slight difference in the input features included in the analysis. For the first outcome (*action to seek help*), data on motivation at pre-study was kept in the feature space, while motivation to seek help at the end of the study was removed. Same applied to the third outcome (*action to seek help at follow-up*). Differently, for the second outcome (*action and motivation to seek help*), all data on motivation at pre- and post-study was removed from the feature space. Implementation details of the three algorithms are summarized next.

### Naïve Bayes classifier (NB)

This algorithm uses Bayes’ rule to predict the probability of an observation to belong to one of the outcome classes given its discriminant function values. This probabilistic model takes continuous and categorical features into account and assumes that they are independent [32]. In the present implementation, after training the algorithm, repeated 10-fold CV was used to evaluate the model’s performance. A step-wise approach was used to select the appropriate number of CV repetitions necessary to achieve accurate and stable performance estimates (50, 100, 150 and 300 CV repetitions).

### Random Forest (RF)

Tree-based methods use recursive binary splitting to stratify the features’ space in smaller, non-overlapping regions used for classification. Trees are easy to interpret but they lack of predictive power, since they tend to overfit the training data. RF can be used to improve prediction accuracy [32,34]. This algorithm requires to tune a set of hyperparameters which control the learning process and are selected (or tuned) by the algorithm to obtain best performance. The following hyperparameters were considered:

▪ Ntree: this value is usually fixed at a computationally reasonable value rather than tuned [32]. Ntree was set to 800. ▪Mtry: a popular value is given by *√p* (where *p* = the number of predictors) [35]. Different search spaces were explored, with Mtry ranging between 1 and 15
▪ Nodesize: different search spaces were explored, with Nodesize ranging between 1 and 20.

Tuning the algorithm and finding the best hyperparameter combination requires to define an optimization algorithm, or search strategy, and evaluation method. We used grid-search with 10-fold CV resampling. To evaluate model performance, nested CV was applied. In this approach, an inner loop tunes the hyperparameters and an outer loop evaluates a wrapped learner, which comprises the classification task, the learner type (RF) and the hyperparameter tuning process. Here a 5-fold CV was applied as outer resample strategy.

### Support Vector Machine (SVM)

The SVM algorithm iteratively identifies a hyperplane that separates labelled classes also in case of non-linear data distributions. SVMs are computationally expensive, but tend to perform very well on a variety of tasks conducted on non-linearly separable classes. Additionally, the algorithm has the advantage to make no assumptions on the features’ distributional properties [32,34]. Similarly to RF, SVM requires hyperparameter tuning. The following hyperparameters were considered:

▪ Kernel: polynomial, radial and sigmoid functions were included in the search space [32].
▪ Degree: the search space was limited to values from 1 to 5 to avoid the risk of overfitting [32].
▪ Cost: it is recommended to tune both cost and gamma (see next point) on the logarithmic scale [36], and a popular search space for cost is from 2^-5^ to 2^15^ [37].
▪ Gamma: this hyperparameter search space was set to the popular range 2^-15^ to 2^3^ [37].

Nested CV was used as previously described for RF. An inner resampling loop (with 10-fold CV) was applied for hyperparameter tuning and an outer loop (with 5-fold CV) for performance evaluation.

### Classification performance measures

The algorithms were evaluated in terms of prediction accuracy on the test set which indicated the overall proportion of cases correctly classified by the model as compared to the observed outcome. However, class imbalance in the sample can negatively impact prediction accuracy, reducing its informativeness as performance measure [38,39]. Matthews Correlation Coefficient (MCC) (yardstick package [40]) was additionally taken into account to evaluate model performance. This coefficient improves over accuracy measures in case of imbalanced datasets [39,41] and can take values from −1 to 1, with 1 indicating perfect prediction, 0 chance prediction and −1 inverse prediction. In addition we computed the confusion matrix (calculateConfusionMatrix(), mlr package). Its output provides the absolute number and the proportion of correct model predictions and misclassifications for each outcome class. For binary outcomes, the confusion matrix allows to estimate model sensitivity (i.e. accurately identifying seekers) and specificity (i.e. accurately identifying non-seekers). In the present study, obtaining high specificity is of particular interest in the context of a hearing mHealth app. Indeed, individuals with HL who are not prone to seek help are the main target population for tailored treatment recommendations and counselling.

#### Feature importance

After identifying the best performing machine learning algorithm, feature importance was considered. Each feature receives a coefficient of importance that indicates its contribution to model performance, regardless of the type of relationship (direction and linearity) between the feature and the outcome. In RF, feature importance is model-dependent and it indicates how much the feature contributes in reducing node impurity. Importance values were retrieved by the function getFeatureImportance() (mlr package) applied on the RF model trained with the tuned hyperparameters. These importance results have the advantage of being inherent to the model and closely tied to its performance [42]. Conversely, there are no model-specific importance metrics available for NB and SVM. For these algorithms, the importance value assigned to each feature corresponds to the area under the ROC (Receiver Operating Characteristic) curve, which is computed from sensitivity and specificity measures [42]. The function varImp() from the caret package [43] was applied on the model trained with the function train() (caret package), after ensuring comparable performance with the same model previously trained in the mlr package.

Information on feature importance was used to identify which features are mostly relevant for telling apart help-seekers and non-help-seekers. No statistical criterion exists to determine which importance value threshold should be used to retrieve relevant features. Hence, three threshold values were inspected (the first 10, 15 and 20 features in their importance ranking order) and evaluated in terms of predictive accuracy and interpretability. Classification accuracy of these three feature sets were assessed on the outcome data obtained at follow-up. For this analysis, the dataset was reduced to *N_2_* = 131 participants who completed the follow-up questionnaire. The important features were fed into the best performing machine learning algorithm from the previous step.

## Results

### Machine learning classification performance

#### Predicting the action to seek help

A summary of the model-specific classification accuracy for the first outcome (*action to seek help*) is provided in Table 5. The three algorithms show similar overall performance accuracy estimates on the test set, correctly classifying about 67% to 70% of the cases in the full dataset (*N_1_* = 185). RF was the best performing algorithm with an accuracy of 70% and an MCC = .28, indicating that the model’s prediction improves to about 20% over chance. By inspecting the confusion matrix, we observed that RF shows high specificity, correctly classifying 91% of the cases belonging to the *no-action* class. The NB classifier (10-fold CV repeated 50 times) showed the best sensitivity compared with the competing algorithms, with 51% cases in the *action* class being correctly classified. RF and NB were selected for feature importance analyses, given the good predictive performance and high specificity of the RF, as well as highest sensitivity of the NB.

**Table 5.** Model-specific overall performance and class-specific classification accuracy for the first outcome measured at study-end.^a^.

| Action to seek help |  |  |  |  |  |  |  |
| --- | --- | --- | --- | --- | --- | --- | --- |
| Model | Hyperparameters |  |  | Overall performance measures |  | Class-specific classification accuracy |  |
| | parameter | space | tuned | test accuracy | MCC | action ($n_{I1}=64$) | no action ($n_{I2}=121$) |
| **NB** |  |  |  |  |  |  |  |
|  |  |  |  | .66 | .26 | .51 | .75 |
| **RF** |  |  |  |  |  |  |  |
|  | ntree | 800 | 800 | .70 | .28 | .31 | .91 |
|  | mtry | [5,15] | 13 |  |  |  |  |
|  | nodesize | [1,5] | 1 |  |  |  |  |
| **SVM** |  |  |  |  |  |  |  |
|  | kernel |  | radial | .67 | .23 | .39 | .82 |
|  | degree | [1,5] | 4 |  |  |  |  |
| | cost | $[2^{-5}, 2^{15}]$ | .01 | | | | |
| | gamma | $[2^{-15}, 2^3]$ | 69.1 | | | | |

<sup>a</sup> Results are based on the full data set ( $N_I=185$ ). For RF and SVM the table additionally shows the hyperparameter search space used in the resampling procedure and the tuned values used for model training and feature importance analysis. The NB model selected was 10-fold CV repeated 50 times. (NB: Naïve Bayes classifier; RF: Random Forest; SVM: Support Vector Machine; MCC: Matthews Correlation Coefficient)

#### Predicting the action and motivation to seek help

Table 6 summarizes the classification performance with respect to the second outcome (*action and motivation to seek help*), which contains three classes (see above). RF provided the highest accuracy with 55% and MCC=.30, as compared to NB (10-fold CV repeated 100 times) and SVM. However, the confusion matrix revealed that none of the three models was able to adequately tell apart individuals within the *no action* class who differ with respect to high versus low motivation. All algorithms can only correctly classify 2 to 25% of cases in the *no action & high motivation* class. Potentially, an improvement in classification accuracy could be achieved with a larger dataset in which the classes are better balanced, and with a more reliable and elaborated measure of the participant’s motivation to seek help. In view of these limitations, the second outcome will not be considered for feature importance analysis.

**Table 6.** Model-specific overall performance and class-specific classification accuracy for the second outcome measured at study-end.^a^.

| Action and motivation to seek help |  |  |  |  |  |  |  |  |
| --- | --- | --- | --- | --- | --- | --- | --- | --- |
| Model | Hyperparameters |  |  | Overall performance measures |  | Class-specific classification accuracy |  |  |
| | parameter | space | tuned | test accuracy | MCC | action<br>($n_{15}=64$) | no action & low motivation<br>($n_{14}=74$) | no action & high motivation<br>($n_{13}=47$) |
| **NB** |  |  |  |  |  |  |  |  |
|  |  |  |  | .50 | .22 | .47 | .67 | .25 |
| **RF** |  |  |  |  |  |  |  |  |
|  | ntree | 800 |  | .55 | .30 | .55 | .80 | .09 |
|  | mtry | [8,10] |  |  |  |  |  |  |
|  | nodesize | [3,15] | 800 |  |  |  |  |  |
| **SVM** |  |  |  |  |  |  |  |  |
|  | kernel |  | sigmoid | .49 | .20 | .53 | .76 | .02 |
|  | degree | [1,5] | 1 |  |  |  |  |  |
| | cost | [$2^{-5}$, $2^{15}$] | 323 | | | | | |
| | gamma | [$2^{-15}$, $2^3$] | $3.05 \times 10^5$ | | | | | |

<sup>a</sup> Results are based on the full data set ( $N_1=185$ ). For RF and SVM the table additionally shows the hyperparameter search space used in the resampling procedure and the tuned values used for model training and feature importance analysis. The NB model selected was 10-fold CV repeated 100 times. (NB: Naïve Bayes classifier; RF: Random Forest; SVM: Support Vector Machine; MCC: Matthews Correlation Coefficient)

### Feature importance

#### Predicting the action and motivation to seek help at follow-up

Feature importance was analysed based on the RF and NB algorithms predicting the first outcome, *action to seek help* at study-end on the full data set (*N_1_* = 185). For both models, features were first ranked in decreasing order according to their importance values. Three sets of features among the most important ones were taken into account for subsequent analysis:

▪ the top-10 features indicated by the two models, resulted in a total of 14 best features;
▪ the top-15 features indicated by the two models, resulted in a total of 19 best features;
▪ the top-20 features indicated by the two models, resulted in a total of 28 best features.

Figure 1 shows all 28 features with their importance ranking values originating from the RF and NB models. Next, the three sets of features were evaluated for their predictive performance and classification accuracy on the reduced dataset of *N_2_* = 131 participants who completed the follow-up questionnaire. NB and RF were trained on the three different feature sets for predicting the action to seek help at follow-up. Results are summarized in Table 7. They show that all feature sets provide good predictive performance and that the NB algorithm outperforms RF, with an overall accuracy ranging between 73% to 75% and an MCC between .43 and .47. Class-specific classification accuracy is comparable between NB and RF, with the action class correctly classified in 52 to 63% of the cases and the no-action class in 82 to 86% of cases. Figure 2 additionally provides an overview of model-specific relative confusion matrices for action to seek help at study end and at follow-up.

**Table 7.** Model-specific overall performance and class-specific classification accuracy for the outcome measured at follow-up.^a^.

| Action to seek help at follow-up |  |  |  |  |  |  |  |
| --- | --- | --- | --- | --- | --- | --- | --- |
|  | Model | Hyperparameters |  | Overall performance measures |  | Class-specific classification accuracy |  |
| | | parameter | space | test accuracy | MCC | action ($n_{21}=52$) | no action ($n_{22}=79$) |
| Top-10 features (total 14) | NB |  |  |  |  |  |  |
|  |  |  |  | .74 | .44 | .55 | .86 |
|  | RF |  |  |  |  |  |  |
|  |  | ntree | 800 | .73 | .43 | .58 | .84 |
|  |  | mtry | [1,4] |  |  |  |  |
|  |  | nodesize | [1,5] |  |  |  |  |
| Top-15 features (total 19) | NB |  |  |  |  |  |  |
|  |  |  |  | .73 | .43 | .58 | .83 |
|  | RF |  |  |  |  |  |  |
|  |  | ntree | 800 | .73 | .41 | .56 | .84 |
|  |  | mtry | [1,10] |  |  |  |  |
|  |  | nodesize | [1,5] |  |  |  |  |
| Top-20 features (total 28) | NB |  |  |  |  |  |  |
|  |  |  |  | .75 | .47 | .63 | .83 |
|  | RF |  |  |  |  |  |  |
|  |  | ntree | 800 | .70 | .36 | .52 | .82 |
|  |  | mtry | [1,5] |  |  |  |  |
|  |  | nodesize | [1,5] |  |  |  |  |

<sup>a</sup> Results are based on the reduced follow-up dataset ( $N_2=131$ ). The table shows the results for three sets of features (10, 15 and 20 most important features for the two models). For NB 10-fold CV repeated 50 times was used. For RF the table additionally shows the hyperparameter search space used in the resampling procedure. In contrast to the previous results (Tables 5 and 6), the tuned hyperparameters were not retrieved, as no importance analysis followed. (NB: Naïve Bayes classifier; RF: Random Forest; SVM: Support Vector Machine; MCC: Matthews Correlation Coefficient)

**Figure 1.**
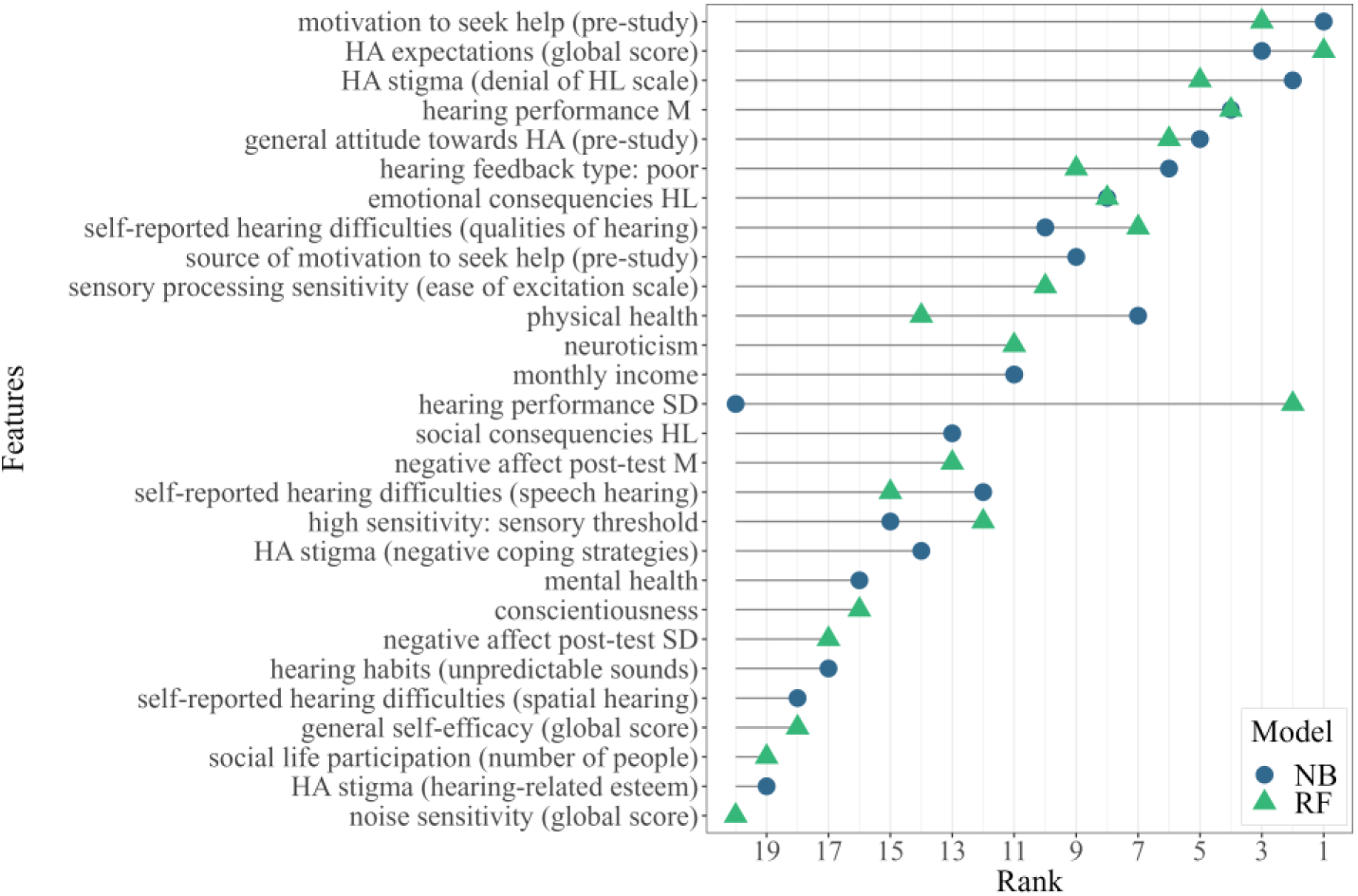
Ranking of the most important features to predict *action to seek help* at study-end. Importance ranking are shown for the 20 most important features for the two models (NB and RF) trained on the full dataset (*N1*=185), as summarized in Table 5. A total of 28 features are arranged on the *y*-axis with respect to their average ranking between the two models. (HL: hearing loss; HA: hearing aids; *M*: mean; *SD*: standard deviation; NB: Naïve Bayes; RF: Random Forest)

**Figure 2.**
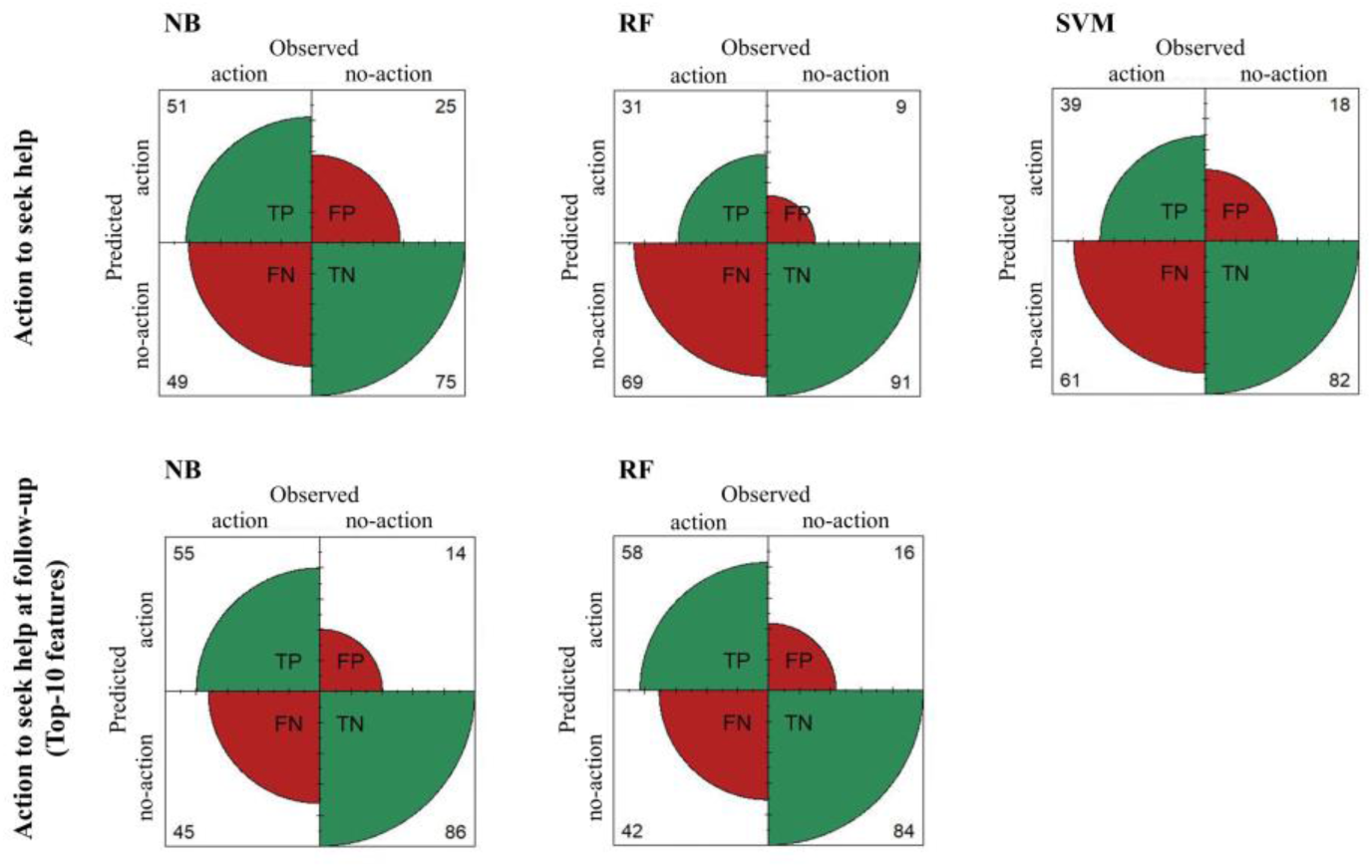
Visualization of the model-specific relative confusion matrices for the outcomes action to seek help, measured at study end (*N1* = 185) and action to seek help at follow-up (*N2* = 131) considering the set of top-10 most important features. Each plot illustrates the class-specific relative frequencies for each observed and predicted class, and provides a more immediate overview of each model’s sensitivity (TP) and specificity (TN). (TP: True positive; TN: True Negative; FP: False Positive; FN: False Negative; NB: Naïve Bayes classifier; RF: Random Forest; SVM: Support Vector Machine)

#### Important features

The following is a brief description of 14 features ranked among the most important (previously named as the top-10 features for the two models) in the prediction of *action to seek help at follow-up*:

▪ *Motivation to seek help* and *source* of this motivation at the beginning of the study;
▪ *Individual’s* attitude and expectations towards HA, including: *general attitude towards HA*; *expectations towards HA*, as measured by the global score of the Expected Consequences of Hearing-Aid Ownership (ECHO) questionnaire [44], which assesses positive and negative expectations towards HA, expected services and costs, and assumptions about change in the personal image in case of HA use; and *stigma towards HA*, as measured by the Denial of Hearing Loss scale of the Attitudes towards Loss of Hearing Questionnaire (ALHQ–3.0) [45], which assesses acceptance of hearing aids and acknowledgement of HL;
▪ *Hearing performance* (mean SRT and its variability as measured by the DTT [26,27]) and *percentage of negative feedback received* (indicating poor performance);
▪ *Perceived* consequences of HL, including: *emotional consequences of HL*, as measured by the corresponding subscale of the Hearing Handicap Inventory (HHIE/A) questionnaire [46]; and *self-reported HL*, as measured by the Qualities of Hearing subscale of the Speech, Spatial and Qualities of Hearing (SSQ) questionnaire [44], which addresses recognition, perceived clarity and naturalness of everyday sounds, as well as listening effort experienced in different hearing contexts;
▪ *High sensory-sensitivity personality*, as assessed through the Ease of Excitation subscale of the High Sensitive Person Scale (HSPS) questionnaire [47], which assesses emotional reactivity to physiological stimuli;
▪ *Reported physical health*, measured by the corresponding items of the Short-Form Health Survey-12 (SF-12) questionnaire [48].

Further features that were assigned lower importance values (among the top-15 features) but might provide further insights toward targeted counselling in a hearing mHealth app are:

▪ *Neuroticism*, which refers to a predisposition of experiencing negative emotions, and *conscientiousness*, which relates to being proactive, organized and methodical (2). These personality traits were assessed through the corresponding items of the NEO Five-Factor Inventory (NEO-FFI) questionnaire [49];
▪ *Monthly income*, which was categorized with three cut-off values (less than 1.500 Euros, between 1.500 and 2.500 Euros, between 2.500 and 4.000 Euros and above 4.000 Euros);
▪ *Social consequences of HL*, as measured by the corresponding subscale of the Hearing Handicap Inventory (HHIE/A) questionnaire [46]; and the number of people included in the individual’s *social networks*, measured by the respective index of the Social Network Index [50].

## Discussion

### Principal Results

The present study contributes to the identification of individuals’ hearing-related, psychological and general health-related features that predict the readiness to seek professional help for hearing loss (HL). Cross-sectional and longitudinal data have been collected in a comprehensive online study. Potential users of a future hearing mHealth app, namely individuals with subjective hearing difficulties, were classified into help-seekers and non-seekers by means of supervised machine learning algorithms. Feature importance analyses helped to derive relevant individual characteristics to be used in a prospective profiling algorithm of help seeking and support the design of a targeted recommendations module in a future app.

#### Which machine learning model can best predict help-seeking and categorize individuals into help-seekers versus non-seekers?

Three machine learning classifiers correctly predicted *action to seek help* at study-end in 66 to 70% of cases, clearly improving over chance prediction. This is a promising result considering the complexity of the prediction outcome. As discussed earlier, several individual factors can influence the decision to pursue hearing health care services and there can be discrepancy between contemplating, planning and taking concrete action [3]. When predicting *action to seek help at follow-up* with the selected important features, the performance of random forest and naïve bayes models improved up to 75%, despite the smaller dataset (*N_2_* = 131). Random forest revealed high accuracy in identifying *no action* both at study-end and at follow-up. Accurate identification of non-seekers is the most relevant performance outcome in an mHealth app to design targeted recommendations. Indeed, the envisioned profiling algorithm should be a system with high specificity that motivates and promotes help-seeking, especially in those cases where users would not spontaneously take action.

#### Which hearing-related and psychological features are most relevant to profile a non-help-seeker?

Hearing performance appears to be one of the most important features to predict help-seeking. The association between degree of hearing loss and help-seeking, as well as with hearing aid (HA) uptake, is well established in literature [4,9,10,12,13]. The present results also highlight – to our knowledge for the first time in the literature – the predictive role of intra-individual fluctuations of hearing performance, emphasizing the need to move beyond the traditional view of hearing as a stable neurosensory process [51]. The implementation of repeated daily measurements of hearing performance provides further insight on the impact of HL on the individual’s everyday life. In line with this, feature importance findings emphasize the relevance of self-reports on the consequences of HL. The assessment should consider self-reported listening effort in different contexts, as well as perceived handicap and potential socio-emotional consequences of HL. Indeed, individuals who report greater negative impact of HL in their life are more prone to seek help and later uptake HA [9,11]. The individuals’ self-awareness of HL can be validated or improved by providing repeated feedback on hearing performance in a hearing mHealth app. As occurred at the follow-up survey, 85% of participants (out of *N_2_* = 131) reported increased awareness for their hearing abilities after receiving repeated feedback during the study. Finally, according to present results and previous findings [10,11,13], investigating stigma, attitude and expectations towards HA informs on individuals’ readiness to seek help, as well as later uptake a HA. Stigma and negative stereotypes related to HA may deter individuals from seeking help and can represent a barrier to HA use [7,12].

Audiological factors emerged as the most important features. Nevertheless, other general health and psychological factors were also relevant in the prediction of help-seeking. In the present study, physical health was an important predictor for help-seeking, although the evidence on this relationship is discordant [10]. While people with better self-reported health were more likely to seek help [4,11], HA uptake was predicted by poor self-reported health [6]. Individuals characterized by high sensitivity to sensory stimuli [22] and emotionally instable personality traits seem to perceive increased psychological discomfort following HL, even in presence of effective HA treatment [2]. Furthermore, it appears that individuals who are proactive and methodical (i.e. with high conscientious trait values) are inclined to plan preventive medical consultations and to show more compliance to treatment recommendations. Finally, income was also listed among the important predictive features. This is in line with evidence suggesting that higher socioeconomic status [9], higher income or pension earnings [3,11] and access to financial support [10] promotes HA uptake.

#### How can feature importance measures inform the design of targeted recommendations for users of a future hearing mHealth app?

Individuals’ attitudes towards HA and self-recognition of HL not only are important predictive features, but could additionally be considered as intervention target to motivate and promote access to hearing care services, where needed. On the one hand, users profiled as decided help-seekers after repeated use of a hearing mHealth app could receive simple and straightforward indications regarding hearing care needed. Those among them who should uptake a hearing device (given their audiological outcome), could benefit from additional information on available hearing-care services and professionals, in order to facilitate faster HA adoption rates. On the other hand, users in need of HA who are profiled as non-seekers should obtain more elaborated, targeted recommendations, which could act as an intervention on modifiable predictive features like self-recognition of HL and attitude towards HA. Non-seekers could receive a more detailed feedback on their hearing performance to improve awareness on their hearing deficit. To promote positive attitude towards HA, information on the large range of devices available, as well as example successful peer cases, could be provided. Knowledge on accessible financial support for HA by insurance companies could additionally promote HA uptake, given the predictive role of income. Furthermore, an implemented HA simulator in a hearing mHealth app could offer possibilities to experience improved listening conditions and promote positive expectations towards a HA. Elaborated information on HA technologies, like the benefits of noise control and noise reduction algorithms, could promote help-seeking by fostering knowledge in individuals who are more sensitive to environmental noise (high sensory sensitivity trait).

The effectiveness of such recommendations could be increased through targeting or tailoring communication. Targeted messages are designed for a specific population segment, while tailored communication is individualized to the person and was shown to be the most effective in promoting health behaviour changes [52]. Indeed, messages that are congruent to the personality traits of the audience are more positively evaluated, persuasive and interesting [53]. The predictive role of personality traits such as neuroticism and conscientiousness for help-seeking behaviour can be considered for efficient communication, both in the context of a hearing mHealth app and in clinical counselling. Individuals with high neuroticism trait are more susceptible to perceived disease [54] and are drawn to action by motives of safety and security [53]. Therefore, they might be more persuaded from recommendations that highlight the positive effects of HA and of an early intervention. Individuals characterized by low conscientious personality require more support to improve adherence to treatment recommendations [55], and would need more guidance in planning the subsequent steps towards help-seeking. For example, a mHealth app would prompt these individuals regularly and remind them on the next step to be taken.

### Limitations and future directions

The predictive performance of the machine learning classifiers could be improved in future studies with a larger dataset and more balanced classes. Classification accuracy could be further improved by including additional objective measures to complement participant self-reports. Continuous psychophysiological measurements (e.g. heart rate variability) could be included as further predictive features. This information could complement the longitudinal assessment of affect and better characterise potential changes in arousal before and after the completion of the auditory measurements. Note that multicollinearity as a potential statistical limitation was ruled out (the correlation plots are available in the Multimedia Appendix 3). Finally, future studies might also benefit from longer follow-up to properly capture those individuals who took more time to take action to seek help. In the present study, measuring help-seeking two months after the end of the study provided a more valid measure of participants’ behaviour. For example, of the participants who were categorized as non-seekers at study-end, seven reported having made an appointment with a hearing professional at follow-up and 12 were planning to do so in the near future.

## Conclusions

This research provides initial knowledge towards a selection of tests and questionnaires to be included in an individual profiling algorithm to predict hearing help-seeking in persons with self-reported hearing difficulties, in the context of a hearing mHealth app. Complementing the audiological assessments with such a profiling algorithm will enable the mHealth app to capture a comprehensive picture of the user and deliver targeted and efficient treatment recommendations depending on those profiles. The benefits of a more comprehensive assessment of the individual with HL might also extend to other applications within a hearing mHealth app. Future studies might explore potential relationships between psychological traits and, among others, HA fitting preferences and endurance in the fine-tuning process towards an optimal aiding solution, openness to try new, elaborated technical solutions, preference for particular app usability features. To conclude, a hearing mHealth app as an easily accessible and affordable mobile diagnostic tool could facilitate faster access to hearing-care services and subsequent earlier intervention, where needed, to pursue the long-term goal of achieving “hearing for all”.

## Supporting information

Multimedia appendices 1, 2 and 3

## Data Availability

The data is available upon request to the corresponding authors. Analysis scripts are available on Zenodo.

https://doi.org/10.5281/zenodo.7635920

## Acknowledgements

This research has been funded by the Deutsche Forschungsgemeinschaft (DFG, German Research Foundation) under Germany’s Excellence Strategy – EXC 2177/1 - Project ID 390895286. The study protocol was approved by the Research Ethics Committee of the Carl von Ossietzky Universität Oldenburg (08.09.2021, Drs.EK/2020/020-01). We thank all the participants who took part in this study and the Universities that helped us with recruitment: Universität Oldenburg, Gasthörstudium; Goethe-Universität Frankfurt am Main, Universität des 3. Lebensalters; Freie Universität Berlin, GasthörerCard Programm; University of Köln, Gasthörer-und Seniorenstudium; University of Kassel, Gasthörendenprogramm; University of Bielefeld, Studieren ab 50.

## Data availability

Analyses scripts are available on Zenodo [56]. The data is available upon request to the corresponding authors. A preprint of the manuscript was published on medrxiv.org in February 2023 and revised in July 2023. Preliminary results of this research were presented on symposia and conferences of the “Hearing4All” Cluster of Excellence, on the VCCA Conference 2022 and the DGPs Conference 2022.

## Author’s contributions

GA, AH and MB conceptualized the study and were involved in protocol development, study design and data analysis. IK was involved in study design and data analysis. BK contributed to study conceptualization and obtained funding. GA was responsible for participants’ recruitment, data collection and wrote the first draft of the manuscript. All authors reviewed and edited the manuscript and approved the final version of the manuscript.

## Abbreviations

CV: cross-validation
HA: hearing aids
HL: hearing loss
MCC: matthews correlation coefficient
mHealth: mobile health
NA: not available
NB: naïve bayes
RF: random forest
SNR: signal-to-noise ratio
SRT: speech recognition threshold SVM: support vector machine

