## Supplementary material for "Predicting hearing help-seeking: What features are important for a profiling module within a hearing mHealth application?": Multimedia appendices 1, 2 and 3

### Multimedia appendix 1 – Daily assessments

#### Affect questionnaire

The affect questionnaire included 14 items in line with the Circumplex Model of Affect (Russel, 1980). Eight items were related to negative affect and six to positive affect. The items were displayed in a randomized order at each presentation and respondents had to indicate on a seven-point scale how much the specified mood applied to them (1 = does not apply; 7 = applies fully). For each of the English affective states retrieved from the Circumplex Model of Affect (Russel, 1980), two German synonyms were selected with the help of native German speakers.

| | English item (Circumplex Model) | German item-pair implemented | |
|---|---|---|---|
| Negative affect items | nervous | angespannt | nervös |
| | sad | traurig | bekümmert |
| | upset | ärgerlich | entrüstet |
| | stressed<br>(item not present in the model) | gestresst | gereizt |
| Positive affect items | excited | begeistert | hocherfreut |
| | happy | fröhlich | glücklich |
| | calm | entspannt | gelassen |

#### Hearing test

Digit Triplets Test (DTT) – provided by the Hörzentrum Oldenburg gGmbH and publicly available under https://www.hz-ol.de/en/ztt.html

Further details on the test:
The speech material of the DTT is represented by random combinations of three digits (eg. "two six five") spoken by a female voice; all numbers between 0 and 9 are included, except from the disyllabic digit 7. The noise of the DTT consists of 30-time superimposition of speech material. Each hearing-test measurement consists of several presentations of spoken digits in noise, that the participant is asked to recognise and repeat on a number pad, for a total testing time of approximately three minutes. The test starts at a +4 dB SNR, so that the speech can be easily recognized by most participants. The SNR is then adjusted at every trial with a one-up one-down adaptive procedure with 2 dB step-size until the individual's SRT can be determined: after each trial, the noise increases if the participant's response is correct, while decreases if it is not. If a maximum value of +10 dB SNR is reached, the test is aborted and the result is categorized as NA. The measurement test error is of 0.7 dB. Participants were invited to familiarize with the test before starting the longitudinal assessment. They were instructed to perform the test with the use of their personal headphones, but three participants reported technical difficulties with their headphones and used loudspeakers instead.

| | | |
|---|---|---|
| Performance categorization and feedback | SRT < -7.1 d B SNR | Good performance (green feedback) |
| | -7.1 >= SRT < -5.1 dB SNR | Intermediate performance (yellow feedback) |
| | SRT >= -5.1 dB SNR | Poor performance (red feedback) |

### Multimedia appendix 2 – Hearing test feedback

Fk. VI | Department für Psychologie
Psychologische Methodenlehre und Statistik

Navigation: [...] > Psychologie > Abteilungen > Psychologische Methodenlehre und Statistik > Testergebnis1

DEPARTMENT OF PSYCHOLOGY
CARL VON OSSIETZKY UNIVERSITY OLDENBURG


Kontakt

Leitung

Prof. Dr. Andrea Hildebrandt
+49 (0)441 798-4629

A07 0-062

Sekretariat

Sandra Marienberg
+49 (0)441 798 -5523

A07 0-035

Anschrift

Carl von Ossietzky Universität Oldenburg
Fakultät VI - Medizin und Gesundheitswissenschaften
Abt. Psychologische Methodenlehre und Statistik
Dep. für Psychologie


#### Hörtestergebnis

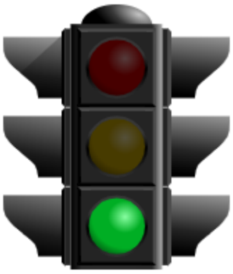

Ihr Hörvermögen bei diesem Hörtest ist normal. Dieser Hörtest kann natürlich nicht alle Aspekte Ihres Hörvermögens abdecken und ersetzt keine medizinische Diagnose. Wenn Sie trotzdem das Gefühl haben, schlecht zu hören, dann sollten Sie einen Hals-Nasen-Ohren-Arzt oder Hörgeräteakustiker konsultieren.

Das Verfahren ersetzt keine medizinische Diagnose, prüft aber das Hörvermögen in einer alltäglichen Situation und kann daher Hinweise geben, wie es um Ihr Gehör bestellt ist.

#### Hörtestergebnis

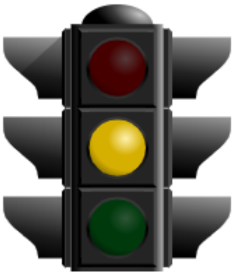

Bei diesem Hörtest verstehen die meisten Menschen etwas besser als Sie. Dieser Hörtest kann natürlich nicht alle Aspekte Ihres Hörvermögens abdecken und ersetzt keine medizinische Diagnose. Falls Sie eine weitergehende Beurteilung Ihres Hörvermögens wünschen, können Sie dafür einen Hals-Nasen-Ohren-Arzt oder Hörgeräteakustiker konsultieren.

Das Verfahren ersetzt keine medizinische Diagnose, prüft aber das Hörvermögen in einer alltäglichen Situation und kann daher Hinweise geben, wie es um Ihr Gehör bestellt ist.

#### Hörtestergebnis

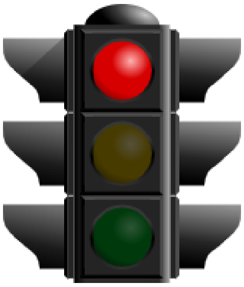

Bei diesem Hörtest verstehen die meisten Menschen deutlich besser als Sie. Dieser Hörtest kann natürlich nicht alle Aspekte Ihres Hörvermögens abdecken und ersetzt keine medizinische Diagnose. Für eine weitergehende Beurteilung Ihres Hörvermögens können Sie einen Hals-Nasen-Ohren-Arzt oder Hörgeräteakustiker konsultieren.

Das Verfahren ersetzt keine medizinische Diagnose, prüft aber das Hörvermögen in einer alltäglichen Situation und kann daher Hinweise geben, wie es um Ihr Gehör bestellt ist.

### Multimedia appendix 3 – Correlation plots

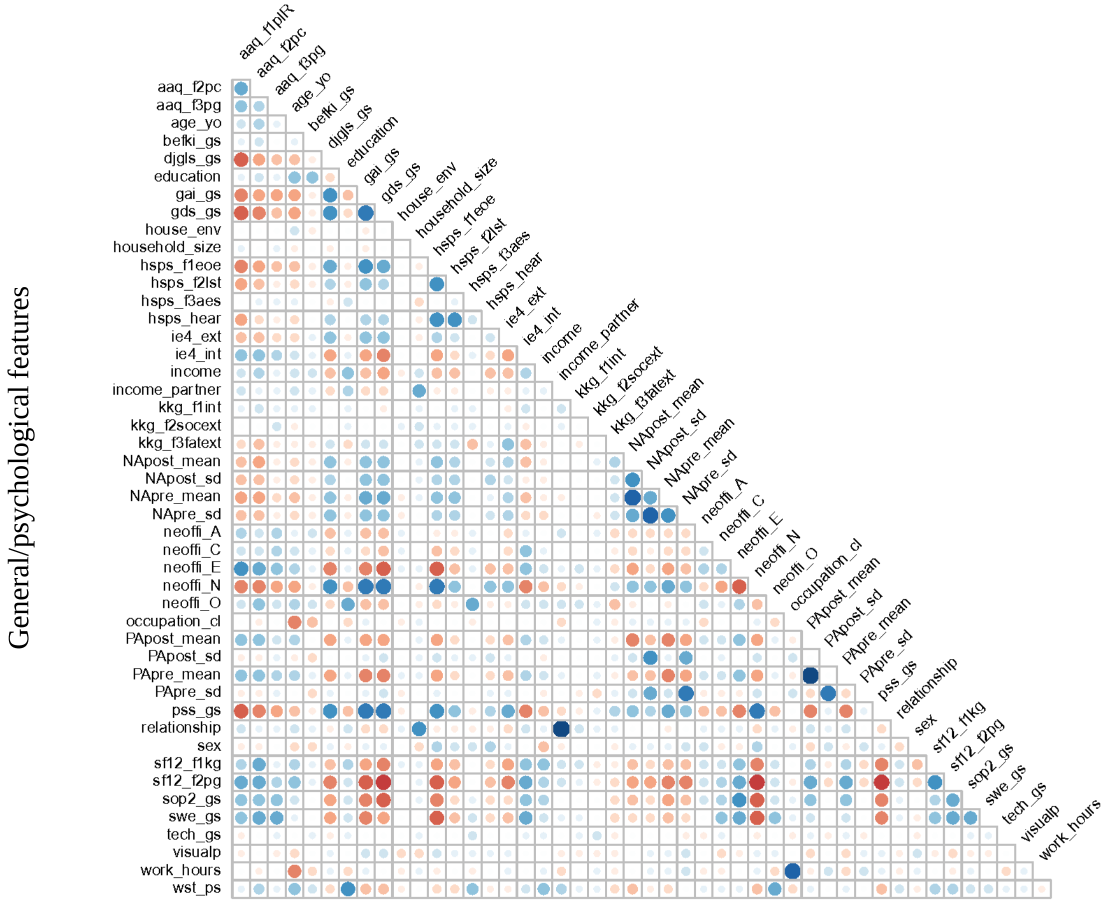


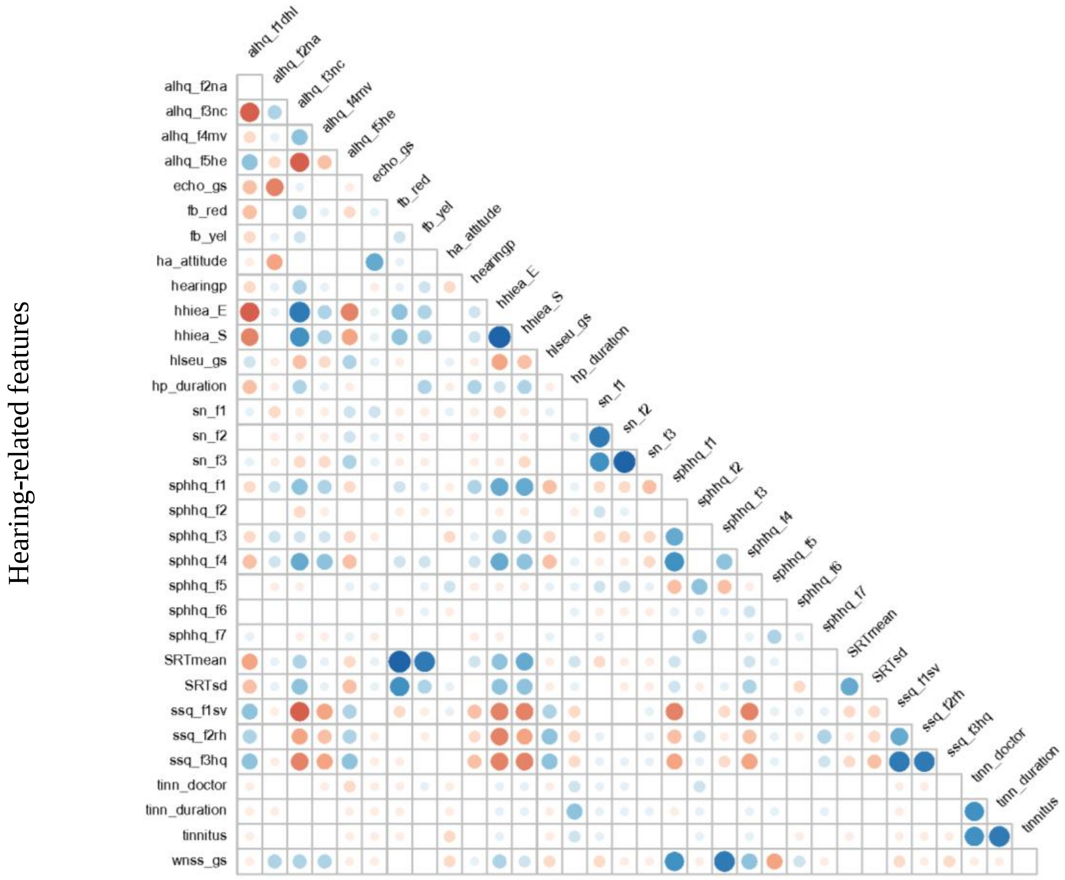
